# Incorrect Versus Correct Mask Utilization in Public Places

**DOI:** 10.1101/2023.07.10.23292470

**Authors:** Thomas F Heston

## Abstract

Mask usage was mandated by public health authorities globally to decrease the spread of COVID-19. These recommendations were based on data showing that N95 masks and possibly surgical masks, when worn tight against the face, help slow the transmission of the SARS-CoV-2 virus. However, cloth and loose-fitting surgical masks are greatly inferior.

**Methods:** Mask use by a random observation of 100 people in public indoor facilities was recorded and statistically analyzed.

**Results:** Out of 100 people wearing a mask, 37 wore a cloth mask. Another 36 people wore a loosely applied surgical mask. Only 27 people wore a surgical mask that covered the nose and mouth and was applied firmly against the face at its margins. There were no people seen wearing an N95 mask. Overall, people were about 70% more likely to wear a surgical mask than a cloth mask (63 vs 37, p < 0.05). Of those wearing a surgical mask, more people wore it loosely than properly (36 to 27, p=0.17). Overall, people were more likely to wear a cloth mask or improperly applied surgical mask than a properly fitted one (73 vs 27, p < 0.001).

**Conclusion:** In public settings, using cloth or loose-fitting surgical masks was almost 3 times more common than adequately using a tight-fitting surgical mask. Out of the 100 people observed, none wore an N95 respirator mask.

## Introduction

Since the SARS-CoV-2 virus was first reported in December 2019, mask mandates were implemented globally to reduce virus transmission and decrease COVID-19, with estimates of up to 95% of the global population living in countries with mask mandates by the summer of 2020 (1). As late as the summer of 2022, mask mandates were still being implemented at schools, some public facilities, and healthcare settings (2,3).

The type of face mask, however, makes a significant difference in transmission rates. One study of 534 participants found that N95 respirators were twice as effective as surgical masks in reducing SARS-CoV-2 virus transmission (4). N95 respirator masks have also been shown to be superior to cloth masks and face shields (5). Studies specifically looking at cloth masks compared with surgical masks in SARS-CoV-2 transmission are sparse. But when looking at the transmission of other virus-like illnesses, surgical masks are about 6 times more effective than cloth masks (6). Some analyses show that cloth masks have no value in reducing respiratory viral disease transmission and are of no benefit in reducing SARS-CoV-2 transmission (7).

This study examined the use of masks in public places in the US. The type of mask was determined, along with how those masks were applied to the face. By evaluating the real-world use of masks, as opposed to laboratory tests or theoretical models, public health officials can create better responses to respiratory pandemics.

## Methods

Mask use by adults in public places was observed in an anonymous fashion. Mask usage was categorized into the following 4 categories: cloth mask, loose fitting surgical mask, properly applied surgical mask, and N95 respirator mask. A surgical mask was deemed loose fitting if the top was below the nose or if large open gaps were around the nose. Those wearing a loose surgical mask, or a cloth mask, were combined into a single category described as incorrect mask usage. Those wearing a tight surgical mask or an N95 respirator mask were combined into a category described as correct mask usage.

Descriptive and inferential statistical analyses were performed by SPSS (8). Statistical fragility was determined using the Unit Fragility Index, the Fragility Quotient, and the Percent Fragility Index (9–11). This research was determined by the University of Washington Human Subjects Division not to involve human subjects, as defined by federal and state regulations (STUDY00018288).

## Results

There were 100 adults in public places observed, and mask use was recorded. Of these, 37 wore a cloth mask, 36 wore a loose surgical mask, and 27 wore a tight surgical mask.

Nobody was observed wearing an N95 respirator mask. A Chi-Square Test of Independence found no significant difference in the use of the cloth versus loose surgical versus tight surgical mask types (Table 1). Those wearing a surgical mask were just as likely to wear it loose as tight (p = 0.172 by Chi-Square).

**Table 1.** Mask use by type

| Mask Type * | n |
| --- | --- |
| Cloth | 37 |
| Loose Surgical | 36 |
| Tight Surgical | 27 |
| N95 Respirator | 0 |

\* $p = 0.255$ by Chi-Square Test of Independence

When mask use was categorized as incorrect (cloth or loose surgical) and compared with correct (tight surgical or N95 respirator), significantly more people were found to use masks incorrectly (73 vs 27). Overall, people were almost three times more likely to wear a cloth mask or wear a surgical mask inappropriately (OR 2.7, p < 0.001) than wear a surgical mask correctly (OR 2.7, p < 0.001, Table 2). The Unit Fragility Index (17), the Fragility Quotient (0.085), and the Percent Fragility Index (23%) all are consistent with high statistical robustness when comparing incorrect with correct mask usage.

**Table 2.** Incorrect mask use was much more common than correct mask use.

| Mask Use * | n |
| --- | --- |
| Incorrect | 73 |
| Correct | 27 |

*\* $p < 0.0001$ by Chi-Square Test of Independence*

## Discussion

There continues to be controversy over whether or not masks have had a meaningful impact on the Covid-19 epidemic (12,13). Part of the reason for this controversy is most likely partly due to the inappropriate use of masks by the general public. While it is well known that inappropriate mask use is common, our findings provide essential information by quantifying the rate of inappropriate mask use.

For masks to be effective, the mask’s seal to the face is critical (14). Thus it is reasonable to categorize cloth or loose surgical mask use as incorrect. When comparing incorrect with correct use, it was found that incorrect use was nearly three times more common than correct use in public settings.

This study was limited because it only examined 100 adults in public settings. However, the findings are not only statistically significant but also powerfully robust. The measures of fragility utilized a theoretically expected rate of 50/50 when comparing incorrect with correct mask use. However, optimal mask use by the public would be closer to 0% to 10% incorrect use versus 90% to 100% correct use. The findings are even more robust when using this as a baseline expected rate. These findings indicate with high certainty that mask use by the general public was predominantly incorrect.

## Conclusion

Incorrect mask use in public settings was about three times more likely than correct mask use. These findings are highly robust and likely widely applicable.

## Data Availability

All data produced in the present work are contained in the manuscript

